# Multiple-region Gray Matter Atrophy contribute to freezing of gait in Parkinson’s Disease

**DOI:** 10.1101/2022.08.10.22278632

**Authors:** Song Zhang, Jie Huang, Dongzhen Liu, Yating Yin, Hua He, Kejia Hu

## Abstract

**Objective:** The aim of our study was to detect the localization of gray matter atrophy in FOG PD patients compared with nFOG PD patients.

**Methods:** A total of 155 PD patients (110 men and 45 women) were included in the current study. Forty-five patients were classified as FOG and one hundred and ten patients were classified as nFOG. A voxel-based morphometry approach was used to investigate the atrophy area of voxel clusters in the gray matter which is associated with FOG.

**Results:** FOG and nFOG PD patients were not significantly different in gender, average age, educational years, disease duration or UPDRS-part I. Compared with nFOG PD patients, FOG patients showed gray matter atrophy in right Inferior frontal gyrus (opercular part), left Superior frontal gyrus, left Superior temporal gyrus, left Amygdala, left Insula, left Medial superior frontal gyrus and left Medial frontal gyrus (orbital gyrus).

**Conclusion:** Our study identified new gray matter atrophy areas in FOG PD patients compared with nFOG patients.

## Introduction

Parkinson’s Disease is the second most common neurodegenerative disorder [1, 2]. It is a progressive, degenerative disease manifested with motor and nonmotor symptoms[2]. Motor symptoms include bradykinesia, muscular rigidity, rest tremor, and postural and gait impairment (also named as freezing of gait (FOG))[3]. FOG is considered one of the most disabling gait disorders in patients with PD[4]. It is characterized by the failure to initiate gait or a sudden interruption of locomotion[5]. Research shows that FOG reduces life quality and the likelihood of independent living[6], and it is independently linked to reduced health-related quality of life (HRQoL)[7].

In recent years, the neuropathological basis of FOG in PD patients has been studied extensively. Degeneration in various brain areas such as parasagittal frontal areas, left postcentral gyrus and cerebellum, have been reported in PD patients with FOG[8, 9]. However, the brain anatomy changes in early-stage FOG patients have not been accurately described yet. Meanwhile, neuroimaging techniques like magnetic resonance imaging (MRI), single-photon emission computed tomography (SPECT), positron emission tomography (PET) have become useful tools for investigating brain anatomy, etc. MRI provides more useful and more accurate structural and functional information about the cerebral cortex, compared with SPECT and PET. Structural MRI is useful to differentiate PD from secondary and atypical forms of parkinsonism[10]. Furthermore, high-field MRI technology with 3.0 Tesla (T) or higher field strengths, spectacular anatomic delineation may further improve sensitivity of MRI to detect smaller lesions[11].

In recent years, there has been more interest in using voxel-based morphometry (VBM) techniques to observe changes in the gray matter, white matter, and cerebrospinal fluid (CSF) of the brain[12, 13]. VBM is a neuroimaging technique widely used for detecting local changes of brain anatomy[14-19]. There are inconsistent reports with VBM to detect gray matter atrophy in FOG and nFOG PD patients. Some studies conclude that significant gray matter atrophy in the right cerebellum (pyramis, declive), left cerebrum (Brodmann area (BA) 21 and 22) and right cerebrum (BA 10 and 6) in FOG PD patients compared to nFOG PD patients[20]. While other studies show significant gray matter atrophy in the left posterior parietal gyrus, left cuneus, precuneus, lingual gyrus, and posterior cingulate cortex[9, 21]. In other words, the relation between FOG and regional brain atrophy is far from clear. It also illustrates that the lack of reproducibility in VBM studies, which may be explained by difference in VBM methods, parameters, template choice, sample size and characteristics of study subjects[22].

In this study, we aimed to detect the localization of gray matter atrophy between FOG patients and nFOG patients with more accurate information and to determine their potential relationships with clinical measures.

## Materials and Methods

### 2.1 Human subjects

In the present study, 155 PD patients (45 for PD-FOG and 110 for PD-nFOG) who met the UK Bank criteria for PD diagnosis were included in the present study. All patients’ data were downloaded from the database PPMI (https://www.ppmi-info.org/). The disease duration of the patients was defined from the symptom onset. Stages of PD patients were evaluated by Hoehn & Yahr (H&Y), Unified Parkinson’s Disease Rating Scale-I (URDPS-I), URDPS-II, URDPS-III. Demographic and clinical data of the patients were summarized in Table 1.

**Table 1.** Demographic and clinical characteristics of the patients.

|  | FOG | nFOG | P value |
| --- | --- | --- | --- |
| Patients | N = 45 | N = 110 |  |
| Gender (M/F) | 33/12 | 67/43 | 0.142 |
| Average age | 64.271±8.709 | 61.614±8.992 | 0.094 |
| Educational years | 16.222±2.679 | 15.327±2.773 | 0.067 |
| Disease duration | 6.659±7.912 | 6.462±6.815 | 0.876 |
| H&Y | 1.689±0.514 | 1.491±0.520 | 0.033* |
| UPDRS-part I | 5.267±3.726 | 4.673±3.504 | 0.348 |
| UPDRS-part II | 6.867±4.060 | 4.400±3.341 | 0.001* |
| UPDRS-part III | 25.067±10.030 | 19.355±8.311 | <0.001* |
| UPDRS-Total | 37.200±13.900 | 28.427±11.676 | <0.001* |

**Note**--Demographics and disease characteristics are displayed as mean ± standard deviation. H&Y-- Categorical Hoehn & Yahr; UPDRS-- Unified Parkinson's Disease Rating Scale.

### 2.2 Imaging parameters

High-resolution structural MRI data of the brain were collected from a Siemens 1.5T scanner at the Department of Baylor college of medicine. High-resolution structural images were acquired by using three dimensional T1-weighted magnetization-prepared rapid gradient echo (3D T1-MPRAGE). The scanning parameters were as follows: repetition time (TR) = 2300 ms, echo time (TE) = 2.98 ms, matrix size = 256 × 240, 176 1 mm saggital sizes.

### 2.3 Image preprocessing for VBM

To find out the structure abnormalities between FOG patients and nFOG patients, we performed a voxel-based morphometry (VBM) analysis for all structural images using Statistical parametric mapping 8 (SPM8). DICOM files were firstly converted into NIFTI files. All T1-weighted images were segmented into gray matter (GM), white matter (WM) and cerebrospinal fluid (CSF). A high-dimensional DARTEL normalization protocol was applied to the GM maps, and resulting segmented grey matter images were normalized into Montreal Neurological Institute (MNI) template space. The parameter of the normalization is “modulated non-linear only”. Images were visually checked after each step in the pre-processing pipeline. A quality check procedure was executed in the VBM8 toolbox, to identify possible outliers from all subjects in the study groups (no patient was excluded in this step because of outliers). Then, these GM images were smoothed with a Gaussian kernel of 8-mm full width at half maximum. After that, in the “specify second level”, we designed a two-sample t test to compare the gray matter volumes between FOG patients and nFOG patients. Then we eliminated possible edge effects, generated a mask covering voxels with GM densities above 0.3 and then applied it to all the GM images. After that, the GM voxels within the cerebellar mask were extracted for further analyses. The analysis was conducted at the voxel level.

### 2.4 Statistical analysis for VBM

We designed a two-sample t test in SPM8 to compare the cerebral and cerebellar atrophy areas between FOG and nFOG patients. Age and sex were used as covariates in our analysis. Volume differences were presented as voxel clusters in an MNI probability map. In the “Result” interface, we set analysis conditions as follows: P value < 0.001 (uncorrected), cluster size > 40 voxels. Furthermore, we set two groups using totally opposite parameters in the “Result” interface (-1 1 0 0; 1 -1 0 0) to localize the specific atrophy areas in two groups. Last, we marked these atrophy areas with special colors and presented 3-dimensional images using Xjview software (https://www.alivelearn.net/xjview/). The Statistical Package for the Social Sciences, Version 26 was used to compute demographic and clinical characteristics. In the software, we used a two-sample t-test to compare various scores and other data if they had statistical difference. A p value < 0.05 was considered to be statistically significant.

## Results

### 3.1 Demographic and clinical characteristics

Forty-five patients with FOG and one hundred and ten patients without FOG were included in the final analysis. The details of the study are provided in Table 1. No significant differences were observed between patients with FOG and nFOG in gender, average age, educational years, disease duration, UPDRS-part I. On opposite, we found that the two group were different with Hoehn & Yahr, UPDRS-part III and UPDRS-Total scores. As expected, patients with FOG had more severe motor impairments (p = 0.001 for UPDRS-part II; p < 0.001 for UPDRS-part III and total score) and higher H&Y score (p < 0.05).

### 3.2 VBM results

Compared with nFOG patients, we found that the reduced GM volume including right Inferior frontal gyrus (opercular part), left Superior frontal gyrus, left Superior temporal gyrus, left Amygdala, left Insula, left Medial superior frontal gyrus and left Medial frontal gyrus (orbital gyrus) (Table 2, Figure 1 and Figure 2). Instead of setting the conditions as p < 0.05 with FWE correction, all results were significant for p < 0.001 (uncorrected) with cluster thresholding (40 voxels).

**Table 2.** Location and MNI Coordinates of significant clusters of GM atrophy

| Cerebellar Regions | L/R | BA | MNI Coordinates |  |  | Cluster Size | T | Z | P |
| --- | --- | --- | --- | --- | --- | --- | --- | --- | --- |
| (ALL Labels) |  |  | X(mm) | Y(mm) | Z(mm) | (voxels) | value | value | value |
| Inferior frontal gyrus, opercular part | R | 46 | 47 | 18 | 28 | 193 | 4.51 | 4.36 | <0.001 |
| Superior frontal gyrus | L | 6 | -15 | 2 | 55 | 49 | 4.19 | 4.07 | <0.001 |
| Superior temporal gyrus | L | 41 | -60 | -31 | 15 | 173 | 3.88 | 3.79 | <0.001 |
| Amygdala | L | 34 | -17 | 2 | -17 | 97 | 3.87 | 3.77 | <0.001 |
| Insula | L | 13 | -42 | -13 | 15 | 109 | 3.85 | 3.76 | <0.001 |
| Medial superior frontal gyrus | L | 9 | -6 | 47 | 18 | 116 | 3.76 | 3.67 | <0.001 |
| Medial frontal gyrus /orbital gyrus | L | / | -3 | 50 | -5 | 92 | 3.66 | 3.58 | <0.001 |

**Fig. 1.**
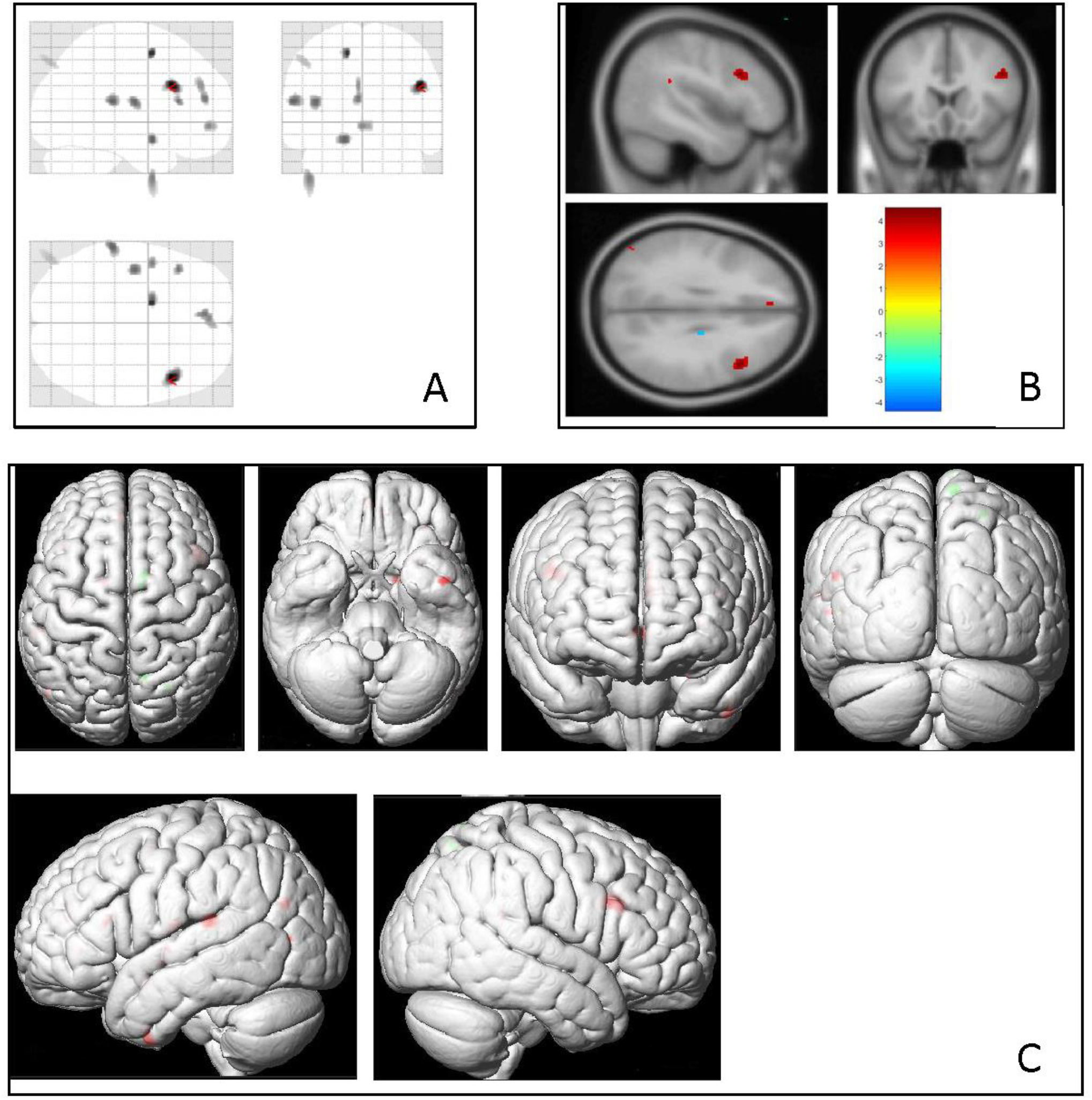
Regions showing atrophy gray matter (GM) volumes in FOG patients compared with nFOG patients, controlling for age and sex (p < 0.001, uncorrected, K > 40 voxels). (A) Regional atrophy GM volumes presented by SPM8. (B) An example of VBM analysis results. In this figure, the red color represents the volumes of the Inferior frontal gyrus (opercular part) in nFOG PD groups that are larger than FOG PD patients. (C) Render view of GM volumes atrophy presented with Xjview.

**Fig. 2.**
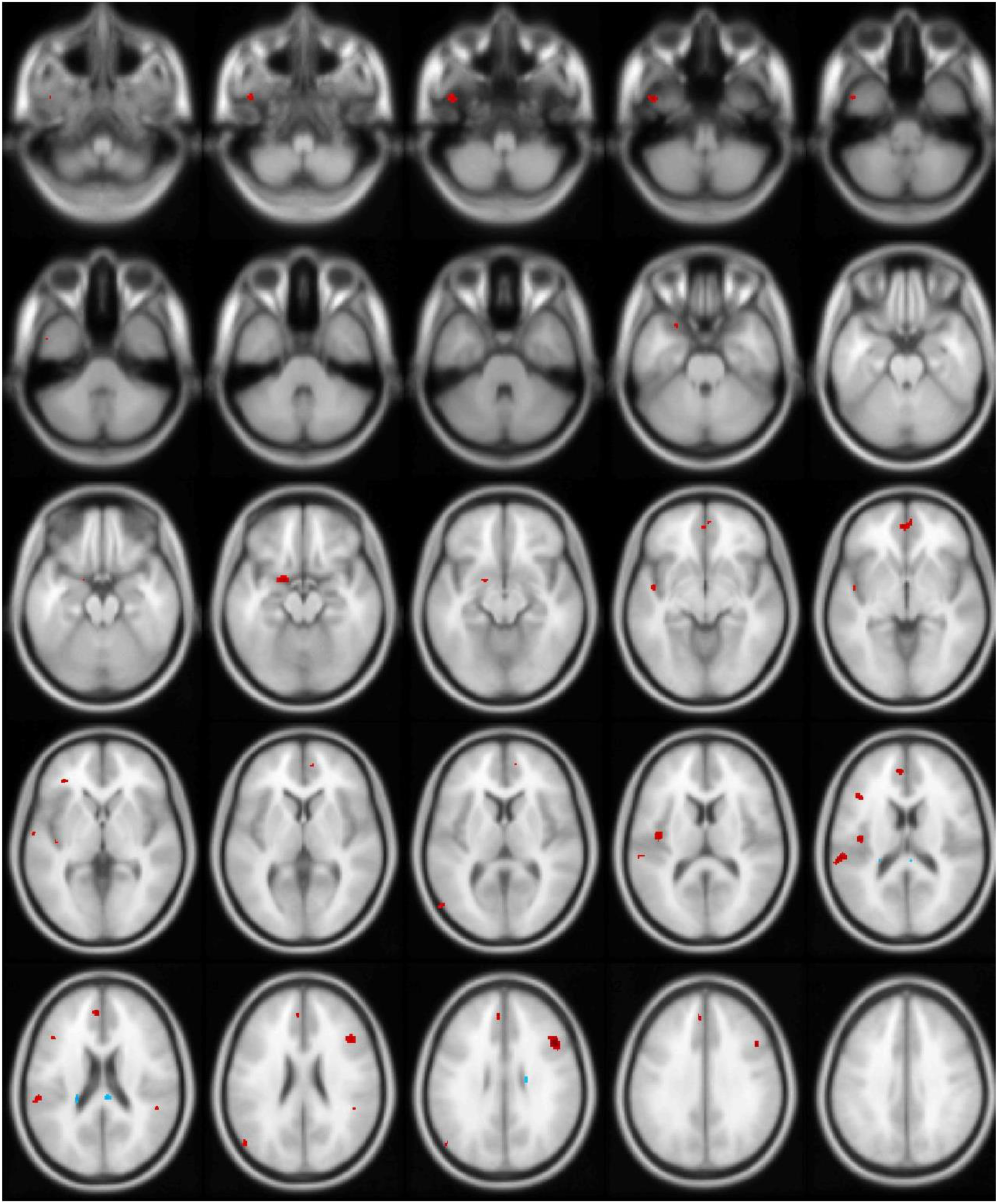
Comparison of gray matter volume between FOG patients and nFOG patients under slice view. Significant changes were indicated by an uncorrected p < 0.001, controlling for age and sex. This figure represents 3-D brain surface rendering graphs, where gray matter volume atrophy was labeled with red color.

## Discussion

The aim of our study was to investigate structural differences in gray matter between PD patients with or without FOG. Our findings revealed that patients with FOG exhibited significant atrophy in left insula, left calcarine fissure, left superior parietal lobule and bilateral superior temporal gyrus compared with nFOG patients. These changes may be the main cause for their clinical symptoms.

So far, the limited results for VBM study of gray matter atrophy in PD patients with FOG seem inconclusive. Menka et al. revealed significant gray matter atrophy in the right cerebellum (pyramis, declive), left cerebrum (Brodmann area (BA) 21 and 22) and right cerebrum (BA 10 and 6)[20]. In their study, patients were grouped by symptoms (with and without FOG) in order to explore the atrophy brain regions in FOG patients and the relation of FOG with recognition. While Florian and colleagues suggested that FOG was associated with focal atrophy in middle frontal gyrus, superior/middle orbital gyrus, superior and inferior parietal lobule (precuneus and supramarginal gyrus, respectively), superior frontal gyrus, and middle temporal gyrus on the right side[23]. And the authors indicated that those clusters which correlated with FOG were well in line with findings from previous studies[9, 24, 25]. Here, we identify, for the first time, FOG patients exhibit atrophy in right Inferior frontal gyrus (opercular part), left amygdala, left medial superior frontal gyrus and left medial frontal gyrus (orbital gyrus) compared with nFOG patients. Our findings support previous studies reporting gray matter atrophy in PD patients with FOG (compared with nFOG patients).

As expected, we found that FOG patients generally had insula atrophy. However, there were minor differences. Those authors suggested that Frontal Assessment Battery (FAB) scores were associated with clusters of GM loss in insula, while we supposed that GM loss in insula was associated with FOG. There are some possible explanations. One explanation could be that atrophy in the same network nodes may contribute to both executive dysfunction and FOG[23]. Also, another study on brain networks associated with anticipatory postural adjustments in PD patients with FOG suggested that the strength of insula connectivity was negatively associated with the severity of FOG[26]. In other words, FOG patients with insula atrophy might be associated with lower insula connectivity compared with nFOG patients.

Moreover, for the first time, we found right inferior frontal gyrus (opercular part) gray matter atrophy in FOG patients compared with nFOG patients. Wang et al. suggested that PD patients with mild cognitive impairment (PD-MCI) group exhibited increased amplitude of low-frequency fluctuations (ALFF) in the Inferior frontal gyrus (opercular part) and the ALFF in this region was positively correlated with the UPDRS total score[27]. One potential explanation could be that our included FOG patients had long disease duration and may present MCI symptom. Pietracupa and his colleagues[28] concluded that PD-FOG patients exhibited bilateral superior frontal gyrus atrophy using FreeSurfer, while Mi et al.[29] revealed that right superior frontal gyrus presented decreased ALFF and was associated with FOG. In agreement with their results, we found left superior frontal gyrus atrophy in FOG PD patients. Also, compared with nFOG patients, FOG patients exhibited left medial superior frontal gyrus atrophy. Recent resting state fMRI (rs-fMRI) studies showed the increased nodal centrality in the medial superior frontal gyrus[30, 31]. The currently available data indicate that widespread functional disruptions in cortical and subcortical brain structures involving multiple brain networks are responsible for FOG in PD[32]. Although the region may not be directly related to the onset of FOG, the functional impairment may be the potential pathogenesis of FOG. Previous rs-fMRI studies suggested that FOG patients showed increased anti-coupling between the frontoparietal network and left amygdala[33]. In this study, the detected amygdala atrophy indicated the dysfunction of amygdala and may worsen FOG. Further follow-up studies to detect the mechanism of the role of amygdala atrophy in FOG onset. We also found that FOG patients had left superior temporal gyrus atrophy compared with nFOG patients. A recent rs-fMRI study concluded that FOG PD patients showed decreased nodal local efficiency in bilateral superior temporal gyrus[32]. The decreased nodal local efficiency could be a potential explanation. Furthermore, we detected left medial frontal gyrus (orbital gyrus) atrophy in FOG PD patients. In previous studies, there were no reports about association between the atrophy of this region and the onset of PD. Malla and his colleagues suggested that patients with long duration of untreated psychosis (DUP) showed significant gray matter atrophy in medial frontal gyrus (orbital gyrus) compared with short-DUP group[34]. While executive dysfunction and FOG share some cognitive aspects[21], the cognitive impairment in our FOG patients might be related to the atrophy of medial frontal gyrus (orbital gyrus). Future studies are required to further delineate the relationship between the region atrophy and FOG and specific mechanisms whether medial frontal gyrus (orbital gyrus) atrophy can lead to FOG.

Our study has some limitations. First, we did not come to a similar conclusion with previous studies[9, 24, 25]. In other words, our results were in partial agreement with above studies and they varied widely. These previous studies concluded that FOG was associated with focal atrophy in predominantly the right mid frontal and the posterior parietal lobe. Different statistical model and covariates may have accounted for the inconsistent results. Previous studies used age, sex, intracranial volume (ICV) or FAB as covariates. Though each study adjusted for important covariates, these varied widely from one study to another. It is also important to note that there is no consent on MRI scanning session, and this may cause a large variance. Second, we did not further investigate the possible mechanism for the left calcarine fissure atrophy in FOG patients. Though we investigated the significant atrophy brain regions in patients with FOG, we did not know whether atrophy of the calcarine fissure may cause FOG. And there are no studies demonstrating this hypothesis. Novel imaging approaches may be needed to probe the mechanism of the atrophy. Third, we did not explore the correlation between clinical symptoms and the atrophy of brain regions. Specifically, part of the URDPS III, FOG-Q scores and other scales which can present different degrees of seriousness weren’t included in our study. Regression analysis and the risk factor of FOG would be investigated in future study.

## Conclusion

In conclusion, we identified new gray matter atrophy areas, for example, left Superior temporal gyrus, in patients with FOG. VBM can be used to identify these GM volumes atrophy and is more sensitive and less time-consuming. Further studies are needed to investigate the brain atrophy of FOG from a different perspective.

## Data Availability

All data produced are available online at www.ppmi-info.org/data.

http://www.ppmi-info.org/

## Author Contributions

HH and KH contributed to design the study. SZ, JH and DL contributed to acquire and analyze the data. SZ, JH, DL, YY, HH, and KH contributed to interpret the findings and draft the manuscript. SZ, HH, and KH contributed to revise the manuscript. All authors approved the submitted version of the manuscript.

## Acknowledgements

PPMI database (www.ppmi-info.org/data) was our primary source of data in this work. For up to data information, please see www.ppmi-info.org/. We gratefully thank all sponsors of PPMI, including Abbvie, Avid Radiopharmaceuticals, Biogen, Bristol-Myers Squibb, Covance, GE Healthcare, Genetech, GlaxosmithKline, Lilly, Lundbeck, Merck, Meso Scale Discovery, Pfizer, Piramal, Roche, Servier, and UCB.

